# Embryo classification beyond pregnancy: Early prediction of first trimester miscarriage using machine learning

**DOI:** 10.1101/2020.11.24.20237610

**Authors:** Tamar Amitai, Yoav Kan-Tor, Naama Srebnik, Amnon Buxboim

## Abstract

**Objective:** Develop a machine learning classifier for predicting the risk of cleavage-stage embryos to undergo first trimester miscarriage based on time-lapse images of preimplantation development.

**Design:** Retrospective study of a 4-year multi-center cohort of women undergoing intra-cytoplasmatic sperm injection (ICSI). The study included embryos with positive indication of clinical implantation based on gestational sac visualization either with first trimester miscarriage or live birth outcome. Miscarriage was determined based on negative fetal heartbeat indication during the first trimester.

**Setting:** Data were recorded and obtained in hospital setting and research was performed in university setting.

**Patient(s):** Data from 391 women who underwent fresh single or double embryo transfers were included.

**Intervention(s):** None.

**Main Outcome Measure(s):** A minimal subset of six non-redundant morphodynamic features were screened that maintain high prediction capacity. Using this feature subset, XGBoost and Random Forest models were trained following a 100-fold Monte-Carlo cross validation scheme. Feature importance was scored using the SHapley Additive exPlanations (SHAP) methodology. Miscarriage versus live-birth outcome prediction was evaluated using a non-contaminated balanced test set and quantified in terms of the area under the receiver operating characteristic (ROC) curve (AUC), precision-recall curve, positive predictive value (PPV), and confusion matrices.

**Result(s):** Features that account for the distribution of the nucleolus precursor bodies within the small pronucleus and pronuclei dynamics were highly predictive of miscarriage outcome. AUC for miscarriage prediction of validation and test set embryos using both models was 0.68-to-0.69. Clinical utility was tested by setting two classification thresholds accounting for high sensitivity 0.73 with 0.6 specificity and high specificity 0.93 with 0.33 sensitivity.

**Conclusion(s):** We report the development of a decision-support tool for identifying the embryos with high risk of miscarriage. Prioritizing embryos for transfer based on their predicted risk of miscarriage in combination with their predicted implantation potential will improve live-birth rates and shorten time-to-pregnancy.

**Capsule:** The risk of first trimester miscarriage of cleavage stage embryos is predicted with AUC 68% by screening a minimal subset of six non-redundant morpho-dynamic features and training a machine-learning classifier.

## INTRODUCTION

Most miscarriages occur during the first trimester of pregnancy (first 13 weeks), accounting for one out of nine clinical pregnancies and for up to one out of three recognized pregnancies (positive pregnancy test).^1,2^ Unlike second trimester miscarriages and third trimester fetal loses that occur due to multiple reasons (congenital defects, placental problems, cervical insufficiency, and infections),^3,4^ aneuploidy is the main cause of first trimester miscarriages.^5^ Early identification of chromosomal abnormalities that permit normal preimplantation development is performed today via preimplantation genetic testing for aneuploidy (PGT-A). PGT-A is an invasive procedure that involves piercing of the zona pellucida and removal of several trophectoderm cells. Since PGT-A typically requires over 24 hours in clinical settings, blastocysts are often cryopreserved, thus eliminating fresh transfers. In addition, failure of amplification and mosaicism in PGT-A contribute to false negative prediction.^6,7^ Since euploid embryos have a higher potential to implant, the implementation of non-invasive methods for early identification of embryos with high risk of miscarriage has the potential to improve clinical performance.^8^

Machine learning-based decision support tools are increasingly incorporated into the healthcare system^9-11^ and specifically in the assessment of the developmental potential of preimplantation embryos.^12^ Supervised machine learning is the most widely used approach in which retrospective datasets with known outcome (*aka* labels) are employed for training a classification model. Prediction of non-labeled embryos is then performed by identifying patterns that are associated with positive or negative outcome using the trained model. The incorporation of time-lapse incubation systems in IVF clinics provided high quality video files that record the course of preimplantation embryo development. These data facilitated the prediction of the potential of embryos to reach blastulation^13,14^ and to implant within the uterus.^15-17^ Retrospectively labeled datasets of significant size can support the training of convolutional neural networks (CNN) that test the relationships between multiple features by convoluting image pixels. CNN’s were used for performing automated morphokinetric annotation^18,19^ and for predicting implantation.^20-22^ Indeed, we found that CNN classifiers of embryo blastulation and implantation identify dynamic features beyond the discrete morphological elements and morphokinetic events that are broadly used by non-convolutional algorithms.^20^

In IVF treatments, embryos that are selected for transfer according to their predicted implantation potential may possess high risk of miscarriage despite exhibiting normal morphological and morphokinetic profiles. To assess the risk of first trimester miscarriage noninvasively at early stages of preimplantation, we assembled a dataset of 464 embryos consisting of 368 embryos with live birth (LB) outcome and 96 embryos with first trimester miscarriage (MC) outcome obtained from four medical centers. Due to the size of our dataset, we employed XGBoost (XG) and Random Forest (RF) machine-learning models and screened a subset of six non-redundant and predictive morphodynamic features based on the time-lapse images of the embryos. Despite the fact that all embryos had been selected for transfer by the embryologists in the clinics based on the same established markers of embryo quality, we were able to predict MC by day-3 from ICSI fertilization with AUC ∼0.7. The implementation of our machine-learning based decision support tool is expected to improve live-birth rates and shorten time-to-pregnancy by deselecting embryos with high risk of MC for transfer.

## METHODS

### Data Acquisition

#### Patients and embryos

Data from 391 women who underwent fresh single or double embryo transfers were collected from IVF clinics of four medical centers as recently reported(Fig. S1a).^20^ All embryos in the dataset were fertilized via intracytoplasmic sperm injection (ICSI). PGT-A – tested embryos were discarded. A total of 464 positively implanted embryos were included in this study as determined on week-5 by ultrasound imaging of gestational sacs. MC labeling of 96 embryos was determined based on negative indication of fetal heartbeat on week 13 or earlier. The remaining 368 embryos were labeled LB in accordance with a confirmed live birth pregnancy outcome.

#### Labeled video database

Anonymized time-lapse video files were recorded by time-lapse incubators (Embryoscope, VitroLife) and imported together with the associated clinical metadata. Data curation was performed using a PostgreSQL database that we assembled and formulated as reported recently.^20^ In short, the database consisted of a front-end website that supported display, query, and data annotation (Hebrew University IT) and maintained by CHELEM LTD. Morphokinetic annotations and quality assurance were performed by qualified and experienced embryologists.^20^

#### Ethical approval

This research was approved by the Investigation Review Boards of the data-providing medical centers: Hadassah Hebrew University Medical center IRB number HMO 558-14; Kaplan Medical Center IRB 0040-16-KMC; Soroka Medical Center IRB 0328-17-SOR; Rabin Medical Center IRB 0767-15-RMC.

### Feature extraction

#### Extraction of morphological features

Static morphological features are routinely used for evaluating the potential of embryos to implant.^23-26^ The embryos were characterized by their two-cell and four-cell fragmentation percentage and blastomere size symmetry, as annotated in adherence to established criteria.^27^ Additional morphological and geometrical features of the zona pellucida, ooplasm, pronuclei (PN’i) and nucleolus precursor bodies (NPB’s) were semi-automatically extracted per the third frame prior to tPNf (∼one hour) using a custom designed code (Python). The orientation angle of the first cleavage plane was evaluated using the seven z-stack images.^28^ The annotation of all morphological features is summarized in Table-1.

**Table 1:** Summary of all embryo features. PN: Pronucleus. NPB: Nucleolus precursor body.

|  | Features type | Quantity | Selected features |
| --- | --- | --- | --- |
| *Morphology* | Blastomere size symmetry (2C and 4C stage) | 2 |  |
|  | Fragmentation percentage (2C and 4C stage) | 2 |  |
|  | PN'i size features | 7 |  |
|  | PN'i location features | 3 |  |
|  | NPBs number distributions | 4 |  |
| | NPBs spatial distributions within and in between PN'i | 11 | $f_1 = (\max_{i,j} d_{i,j})/R_{pn}$ where $d_{i,j}$ is the distance between NPB $i$ and NPB $j$ within the smaller PN of radius $R_{PN}$ |
|  | 1<sup>st</sup> cleavage plane orientation | 1 |  |
| | Ooplasm and zona pellucida radii | 5 | $f_2 = \text{Ooplasm radius}$ |
| *Morphokinetics* | Morphokinetic events | 9 |  |
|  | Pairwise intervals | 36 |  |
|  | Pairwise time ratios | 36 |  |
| | Distance from linear regression (DLR) | 36 | $f_3 = \text{DLR}_{2,3}$<br>$f_4 = \text{DLR}_{5,6}$ |
| *PN dynamics (Optical flow)* | PN'i step size and velocity | 72 | $f_5 = \text{Abs}(\text{largest step size of larger PN} / \text{largest step size of smaller PN})$ |
|  | Distances between the PN pairs | 18 |  |
| | PN distances from embryo center | 72 | $f_6 = \text{Abs}(\text{distance of larger PN from embryo center minus distance of smaller PN from embryo center at tPNf})$ |

#### Extraction of morphokinetic features

Time-lapse incubators facilitate the annotation of the time points of discrete events. Morphokinetic annotations and quality assurance were performed by qualified and experienced embryologists as we recently reported.^20^ In short, morphokinetic profiles were obtained via majority voting according to established protocols^29,30^ and further validated blindly by an expert embryologist. Morphokinetic annotations included the time of PN appearance (tPNa) and fading (tPNf), and cleavage of N blastomeres (tN, N=2 to 8).^31,32^ We also introduced all 36 pairwise intervals and pairwise ratios between the events.

#### DLR

We calculated the distances from the linear regression (DLR) as follows. Pairs of the morphokinetic features of LB embryos appear to be linearly correlated as demonstrated for t2-t3 events (Fig. S2). These morphokinetic relationships are defined by the linear regression of the morphokinetic scatter plot (R^2^: 0.87). To estimate the association with the averaged morphokinetic profiles of high quality positively-implanted embryos, we calculated the shortest Euclidean distance of each embryo from these linear Regression (Fig. S2).

#### PN dynamics

Male and female PN’i were visible for 17±2.5 hours (average ± STD) between tPNa and tPNf. The trajectories of the PN’I were extracted using Optical Flow as follows. Fifteen non-adjacent pixels were selected within each PN using the starting points frame. The time-lapse consecutive coordinates of each episode relative to the embryo frame of reference were obtained via the Lucas-Kanade model and a 15×15 pixels surroundings.^33^ Both forward (tPNa to tPNf) and reverse (tPNf to tPNa) Optical Flow tracking episodes were performed, consisting of all the time-lapse images in between. The trajectory of each PN was calculated by averaging the coordinates of all 15 tracks.

### Prediction algorithms and data analysis

#### Prediction models

Due to the limited size of the dataset, we employed XGBoost (XG) and Random Forest (RF) models that minimize the risk of overfitting by integrating ensembles of multiple classification processes.^34,35^ In addition, XG and RF are based on clusters of decision trees that are suitable for identifying complex patterns which are derived by nonlinear relationships between the features. The XG model was implemented using XGBoost Python library and the RF model was implemented using the Scikit-learn Python package. We set the number of trees and depth 6 and 1 for the XG model, and 65 and 3 for the RF model. We set 0.25 learning rate for the XG model and the RF splitting criterion was defined based on Gini impurity. Parameter search optimization over the above models was performed using the Hyperopt Python framework package.^36^

#### Validation and testing methodologies

The embryos were divided into a train/validation set and an uncontaminated test set with balanced numbers of MC and LB labels (Fig. S1b). To effectively increase the variation between train set embryos, we performed a 100-fold Monte-Carlo cross validation scheme. Specifically, the classifier was trained on randomly selected 90% of the relevant train-validation set embryos and validated on the remaining 10%. In this scheme, each embryo was selected for validation multiple times, thus increasing the phenotypic variation for MC prediction. MC prediction was performed by averaging the cross-validated model’s weights.

#### Statistical analyses and graphical design

Statistical analyses were performed using NumPy, Scikit-learn, pandas and SciPy Python packages. Figures were generated using Matplotlib and Seaborn Python packages. Manual features were extracted using the Pillow Python package.

## RESULTS

Motivated by the prediction of pregnancy using machine learning, we hypothesized that early pregnancy loss can also be predicted based on time-lapse imaging of preimplantation development. Hence, we assembled a dataset of videos of MC- and LB-labeled embryos that were collected from four IVF clinics that accept patients with a diverse ethnic and racial backgrounds (Eastern and Western European Jews, North African and Middle Eastern Jews, Arabs, and others).^37^ This embryo dataset spans a range of maternal age. Clinics H1 and H3 tend to perform double embryo transfers on day-3 of preimplantation development or earlier whereas single embryo transfers are frequently performed in clinics H3 and H4 in which half of the embryos are transferred on day-5 or later (Fig. S1a). We first compared the morphokinetic profiles of MC and LB embryos, and found indistinguishable profiles, suggesting that the risk of MC cannot be assessed using the standard statistical approaches (Fig. S3a). Similarly, there were no differences between MC and LB embryos in terms of their developmental states and day of transfer (Fig. S3b). We conclude that the ethnic diversity, maternal age distributions and inherent variation in the clinical protocols between the hospitals contribute to the generality of our study. In addition, the overlapping morphokinetic profiles, day of transfer and the developmental states of the transferred embryos are consistent with communal criteria for selecting both MC and LB embryos for transfer, thus minimizing the effects of confounding parameters and satisfying a natural experiment.

### Feature Extraction

To identify visual markers of first trimester miscarriage, we assembled an expansive set of 314 features that were derived from measurable static and dynamic properties of preimplantation embryo development and span a range of length scales and time scales from the NPB to the zona pellucida and from time of PN appearance (tPNa) to time of eight-blastomere cleavage (t8; Table-1 and Methods). The feature set included morphological, morphokinetic and other dynamic features as defined in the Methods section and summarized in Table-1. Morphological features account for specific geometrical properties of the embryo as derived from single frames. Morphokinetic features included the time points of morphokinetic events, their pairwise intervals and pairwise ratios, and the DLR features that quantify an effective distance of an embryo from the average profile of positively implanted embryos.

The remaining features are associated with PN dynamics as evaluated using Optical Flow (Methods). Optical Flow tracking was validated against manual tracking, showing only insignificant disagreement that was upper-bounded by one quarter of PN radius (Fig. 1a). Interestingly, distinctive PN trajectories were observed and classified into one of the following categories based on their end-to-end distance and contour curvature (Fig. 1b): (i) Curved trajectories. (ii) Linear trajectories. (iii) Stationary trajectories. By obtaining the temporal coordinates of the PN’i trajectories with respect to the embryo frame of reference, we defined specific features that were related to PN step size and velocity, the distribution of distances between PN’i, and their distance from embryo center of mass (Table-1).

**Figure 1:**
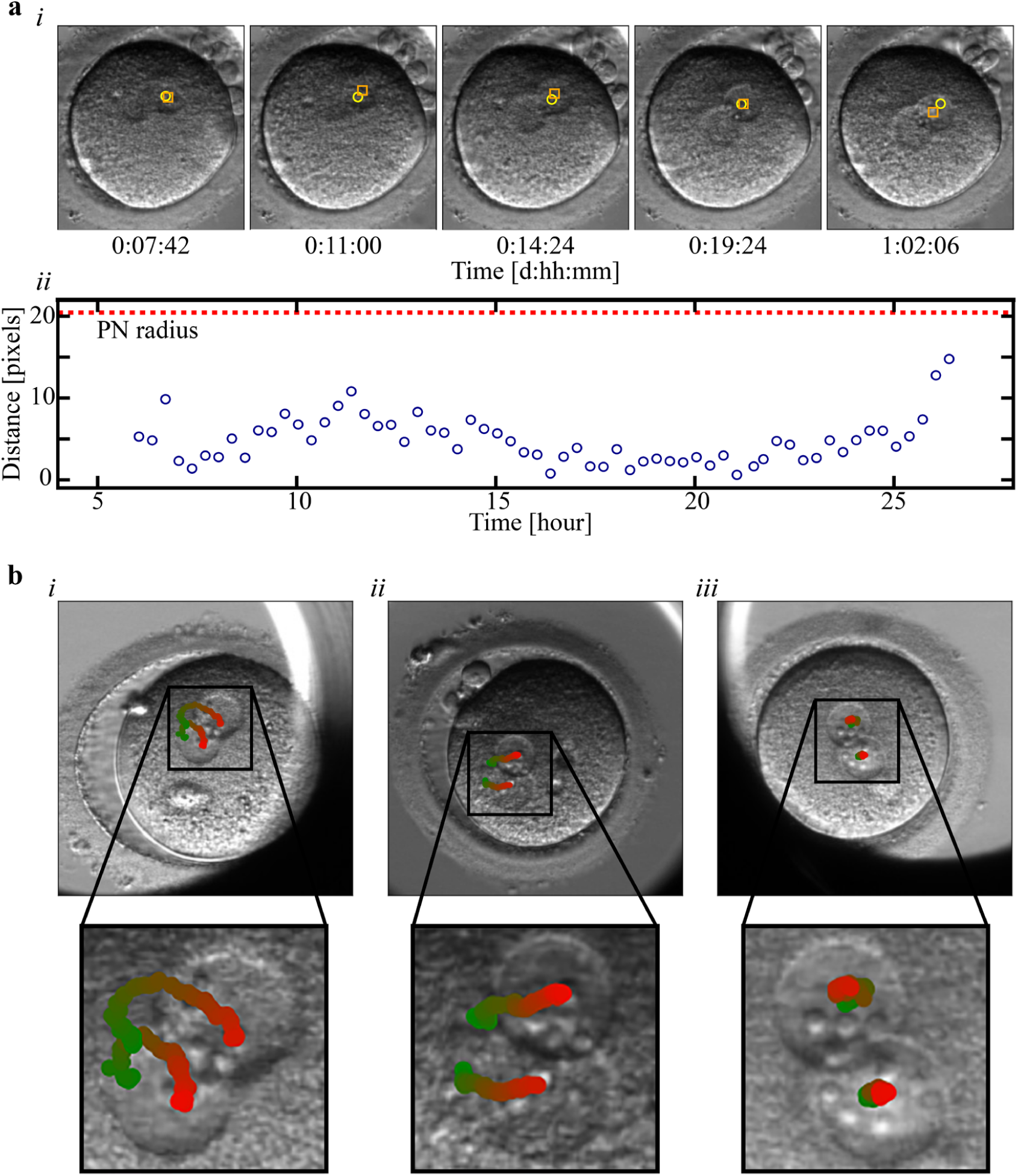
Trajectories of PN dynamics depicted using optical flow. **a**, Validation of the optical flow algorithm: (i) Time-lapse images show a comparison between the dynamic trajectories of a representative PN that was obtained by optical flow (yellow) and via manual tagging of the PN center of mass (orange). (ii) The distance between the locations of the PN as obtained by optical flow versus manual tagging is plotted as a function of time from tPNa to tPNf. The disagreement between automated optical flow and manual tagging is significantly smaller than the relevant length scale set by the PN radius. **b**, Optical flow tracking revealed distinctive types of PN trajectories relative to embryo center of mass: (i) Curved trajectories. (ii) Linear trajectories. (iii) Stationary trajectories. Bottom images show zoom-in of PN trajectories. The temporal appearance of the PN trajectories are color-coded from green (tPNa) to red (tPNf).

### Feature Screening

The breadth of the feature set allowed us to examine a variety of morphodynamic elements and processes. However, the size of the labeled embryo dataset limited the number of training features in order to avoid overfitting. Hence, we carried out feature screening following a stepwise scheme based on statistical distance criteria between MC and LB embryo distributions followed by MC prediction performance using random forest (RF) and XGBoost (XG) models. Statistical distance between MC and LB embryos were calculated by the Kolmogorov-Smirnov (KS) test and the Kullback–Leibler (KL) divergence (Fig. 2a). Four features were distinguished by high KL-entropy and low KS p-value (green symbols): (1) t2-t3 distance from linear regression (DLR2,3; Methods); (2) Maximal distance between the NPBs of the small PN; (3) The difference between the distances of the two PN’i from the embryo center of mass (at tPNf); (4) The maximal distance between pairs of NPBs in the small PN normalized by small PN radius. These features were thus expected to provide high predictive strength.

**Figure 2:**
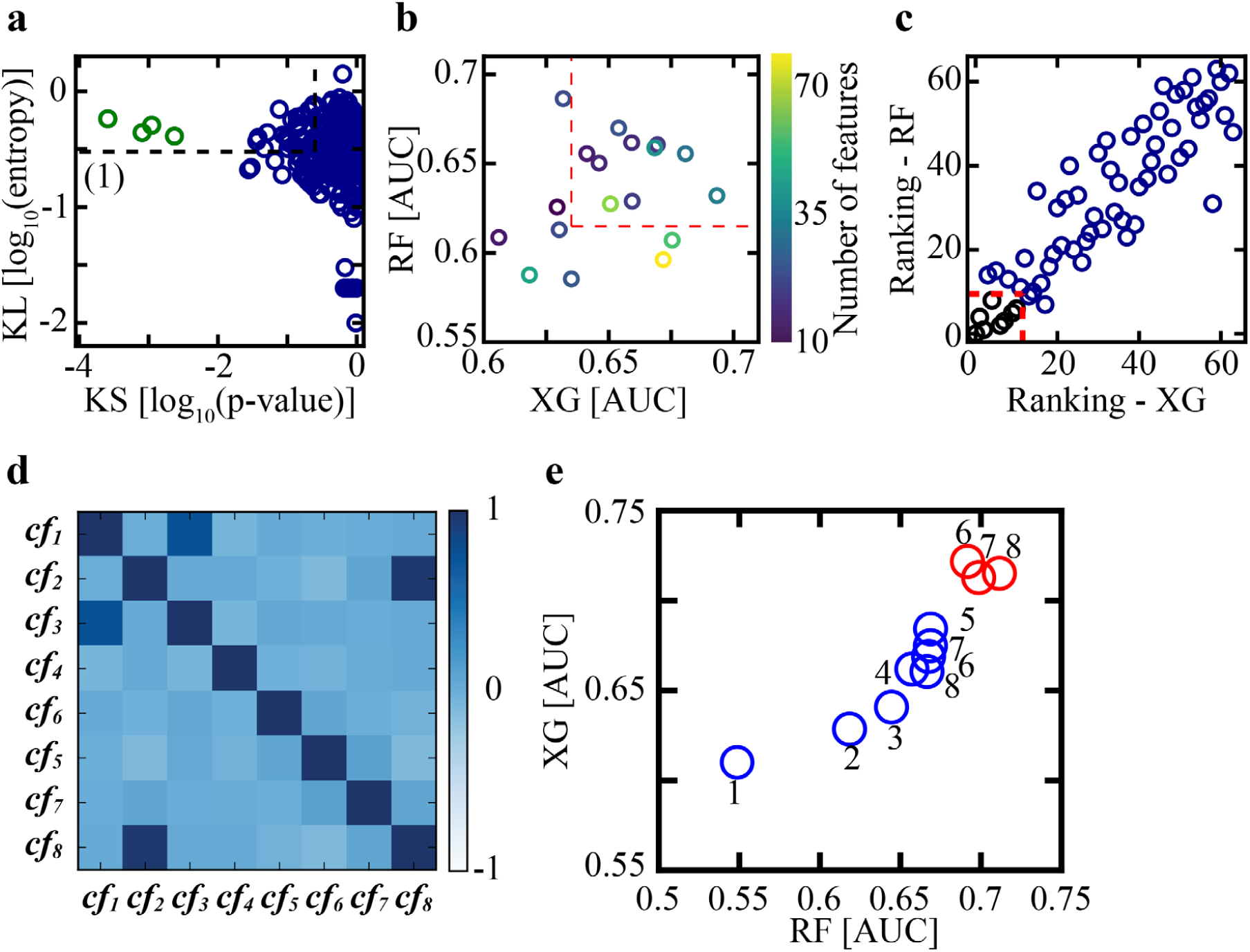
Feature screening. **a**, The statistical features distance (n=314) between their average MC and LB values are quantified using KS test (p-value) and KL divergence (entropy). **b**, RF and XG models were trained on subsets of features with KL entropy above threshold and KS p-value below threshold (dashed black frame in **a**). Using a series of KL and KS threshold values (black frame in **a**), 18 subsets of qualified features were selected and are plotted with respect to their AUC coordinates of MC prediction by RF and XG models. The ten most-predictive subsets consisting of 64 features were selected (red frame). **c**, The selected 64 features were ranked with respect to their XG and RF weights as obtained by 100-fold cross validation within each subset experiment. The top eight ranked features are marked in black (red dashed frame). **d**, Pairwise Pearson correlation matrix of the set of eight features highlights redundancy between cf1 and cf3 and between cf2 and cf8. **e**, Blue symbols: Starting with a set of eight features, XG AUC values increase with the removal of three features as part of stepwise backward feature selection. Further removal of features decreases both RF and XG prediction. Red symbols: Starting with five backward selected features, the addition of one feature via two-step forward feature selection increases both RF and XG AUC values. Addition of subsequent features does not improve prediction. The group of five backward selected features (flue) and one forward selected feature (red) constitute the minimal subset of non-redundant features that retain predictive strength. RF: Random forest. XG: XGBoost. KS: Kolmogorov-Smirnov. KL: Kullback–Leibler.

KS-KL statistical distance criteria estimate the predictive potential of each feature individually. Since our goal is to identify the minimal group of features with high predictive strength, we applied a series of KS p-value and KL entropy thresholds, selected the subset of qualified features with lower KS p-value and higher KL entropy, and assessed their predictive potential by training RF and XG models. Six KS p-value thresholds, 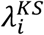, and three KL entropy thresholds, 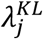, were chosen (Fig. 2a, black frame), thus defining eighteen feature subsets:

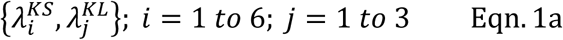

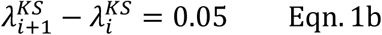

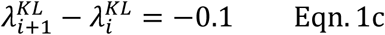

To better assess classification performance by simulating multiple validation sets, we executed a 100-fold Monte-Carlo cross-validation experiment (Methods). The predictive strengths of the selected feature subsets were evaluated by averaging the cross-validated AUC values of the RF and the XG models (Fig. 2b). Next, we selected the ten feature subsets with the highest RF and XG AUC values (Fig. 2b, red frame). In total, these ten subsets included 64 features; a fraction of which were shared by multiple subsets. The features were then ranked by their RF and XG importance scores, as averaged across all relevant Monte-Carlo cross-validation experiments (Fig. 2c). In this manner, we identified the eight top ranked features (Fig. 2c, red frame).

### Selection of a non-redundant feature subset

So far, features were screened based on their individual MC-LB statistical distances and their weighted contributions to RF and XG prediction models. However, redundancy among the eight selected features cannot be excluded. To obtain further insight, we calculated the pairwise Pearson Correlation Matrix between the distributions of their feature values (Fig. 2d). Indeed, feature pairs [cf1, cf3] and [cf2, cf8] were highly correlated. Specifically, the difference between the smallest steps of the two PN’i (cf1) and the difference between the slowest velocities of the two PN’i (cf3) were highly correlated. Likewise, the correlation between the non-normalized (cf2) and the normalized (cf8) maximal distance between NPBs was also high (see Table-1).

To identify the minimal subset of non-redundant features that maintain high predictive strength, we first performed feature selection via backward elimination. Starting with the selected group of eight, each feature was removed in its turn and RF and XG 100-fold Monte-Carlo cross-validation was performed on the remaining subsets of seven features (Methods). The single feature that had the smallest negative effect on RF and XG AUC’s was removed. Backward feature elimination was performed in a stepwise manner, thus identifying a series of subsets with a decreasing number of features from eight-to-one (Fig. 2e, blue symbols). We found that the removal of the first three features did not decrease both RF and XG prediction strengths. However, subsequent removal of the remaining features decreased both XG and RF prediction performance, indicating that the subset of five features represents the minimal collection of non-redundant features with high MC prediction. Consistent with the pairwise Pearson correlation analysis discussed above, cf8, cf2 and cf3 features were indeed discarded in this order (Fig. 2d).

Backward feature selection did not exclude the possibility that reintroducing additional features from the original pool of features that had been screened out wouldn’t improve MC prediction. The goal was to assess the integrated contributions of multiple features rather than testing features one by one. Since testing all combinatorial subsets is computationally not feasible, we employed a two-step forward feature selection scheme. Starting with the subset of five-features, the addition of all feature pair combinations was tested by training RF and XG models as described above. The feature pair that improved prediction the most was identified and the feature with the lower index was arbitrarily selected. In this manner, we performed a five+two, six+two and seven+two forward feature selection cycles thus increasing subset size from five to eight features (Fig. 2e, red symbols). We found that introducing the sixth feature indeed improved the predictive strength of both RF and XG models. However, subsequent addition of the seventh and eighth features failed to increase AUC values. We thus conclude that minimal redundancy and high predictive strength of MC outcome is provided by the final subset of five backward selected features plus one forward selected feature as summarized in Table-1 (right column).

The minimal subset of non-redundant and highly predictive features is described below.

***f***_**1**_ : The maximal distance between NPB pairs within the small PN normalized to PN radius (Fig. S4a):

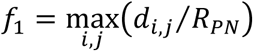

***f***_**2**_ : Radius of the ooplasm.

[Both ***f***_**1**_ and ***f***_**2**_ were evaluated using the central focal plane image that was recorded one hour prior to *PN*_*f*_].

***f***_**3**_ : *DLR*_2,3_ (Fig. S2).

***f***_**4**_ : *DLR*_5,6_.

***f***_**5**_ : The ratio between the maximal step size of the large PN, 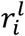, and the small PN, 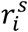, (Fig. S4b):

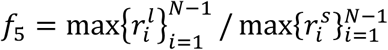

Maximal steps are evaluated across *N* steps performed by the PN’i from *tPN*_*a*_ and *tPN*_*f*_.

***f***_**6**_ : The absolute difference between the distances of the large (*l*_*l*_) and small (*l*_*s*_) PN from zygote center of mass at *PN*_*f*_ (Fig. S4b):

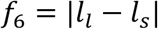

We note that half of the features are associated either with NPB pairwise dynamics (***f***_**1**_) or with PN dynamics (***f***_**5**_ and ***f***_**6**_). The remaining features correspond to ooplasm size (***f***_**2**_) and to cleavage-stage morphokinetic profiles (***f***_**3**_ and ***f***_**4**_).

### Prediction of MC outcome

MC prediction was performed by training an XG model using the selected feature subset *f*_1_ to *f*_6_ (Fig. 3a). The earliest time of MC prediction is set by the time of six blastomere cleavage event as required for feature *f*_6_. To effectively increase the variation between train set embryos, we performed a 100-fold Monte-Carlo cross validation scheme (Methods). As expected, the area under the receiver operating characteristic (ROC) curve (AUC) of train set embryos was relatively high, indicative of moderate overfitting, which is consistent with the size of the available dataset. Nevertheless, AUC of 68-69% was robustly obtained both on the validation set and the test set embryos. Importantly, comparable prediction performance was also obtained both by an RF model and by an integrated XG-RF classifier, thus increasing confidence in the utility of this feature subset for MC prediction (Fig. S5).

**Figure 3:**
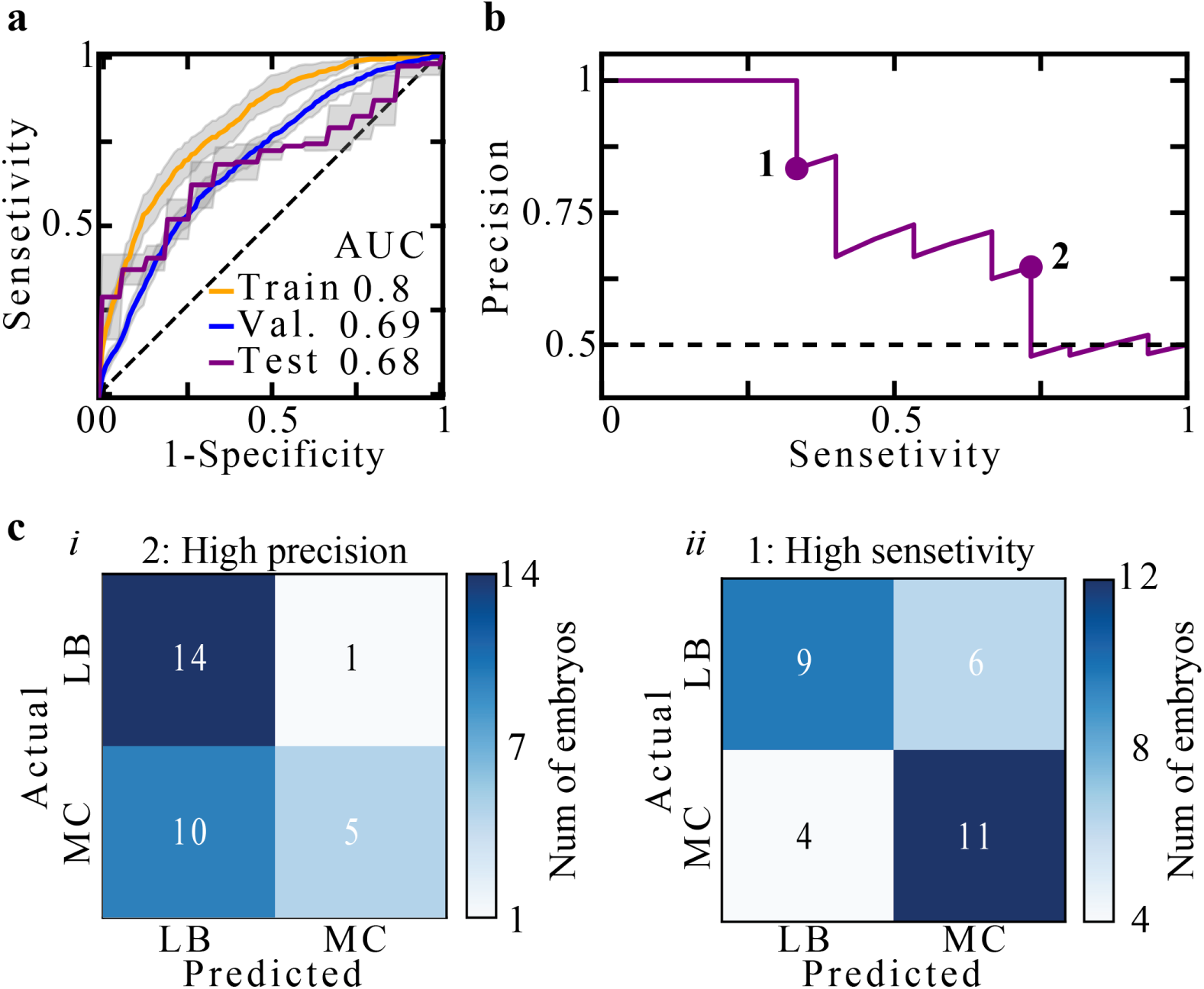
Embryo MC prediction at early preimplantation stages. **a**, Train, validation and test set ROC curves and AUC values of MC prediction using an XG model. Sensitivity confidence intervals, as derived based on a 100-fold Monte Carlo cross-validation scheme, are depicted by grey margins. **b**, A precision-sensitivity curve of test set embryo is plotted. **c**, Confusion matrices of two binary classifiers that favour (i) precision and (ii) sensitivity are evaluated for test set embryos. The binary classifiers were defined by setting the threshold values depicted in b’. ROC: Receiver operating characteristic. AUC: Area under the ROC curve. RF: Random forest. XG: XGBoost.

To demonstrate clinical utility and delineate the trade-off between precision (positive predictive value; PPV) and sensitivity (recall) in relation with improving livebirth rates and shortening time-to-pregnancy, we illustrate two extreme scenarios (Fig. 3b). We first consider a women patient having only a small number of high quality embryos with a predicted high potential of LB outcome. Due to their small number, embryos with both moderate and high predicted LB score should not be discarded by setting a high classification threshold for MC prediction (equivalent to setting a low threshold for LB prediction; Fig. 3c-i). The tradeoff of setting a high MC threshold is the inclusion of embryos with moderate predicted risk of MC outcome. In the opposite case, we consider a woman patient with a large number of embryos with of a high predicted LB potential. Here, the clinicians have the benefit of discarding all embryos with a predicted moderate-to-high risk of MC outcome while also discarding embryos with moderate predicted LB potential by setting a relatively low threshold for MC prediction (equivalent to setting a high threshold for LB prediction; Fig. 3c-ii). Precision is favored in first scenario, which supports increasing livebirth rates at the expense of not shortening time-to-pregnancy due to MC. In the second scenario the clinicians can sustain losing high-quality embryos in order to shorten time-to-pregnancy by lowering the risk of MC.

### Feature importance

PN and NPB morphologies were previously associated with aneuploidy.^38-43^ Here we find that the morpho-dynamic properties as characterized by the trajectories of the PN’i and the NPB’s differentiate between the potential of MC versus LB outcomes. The maximal distance between NPB pairs within the small PN (*f*_1_) is greater in MC embryos than in LB embryos (Fig. 4a; p-val 0.001). In addition, MC outcome is associated with a larger ratio between the maximal step sizes of the large versus the small PN’i (*f*_5_) and a shorter difference between PN’i locations with respect to zygote center of mass (*f*_6_). Morphokinetically, *DLR*_2,3_ distances (*f*_3_) are longer in MC embryos whereas *DLR*_5,6_ distances (*f*_4_) are shorter. This indicates that the potential of LB is more closely associated with the averaged profile of positively-implanted embryos with respect to *t*2 − *t*3 correlations (p-val < 0.01) and that the potential of MC is more closely associated with the averaged profile of positively implanted embryos with respect to *t*5 − *t*6 correlation (p-val < 0.04). Lastly, we find that ooplasm radius (*f*_2_) of MC embryos is slightly larger than of LB embryos (Fig. 4a; p-val < 0.01).

**Figure 4:**
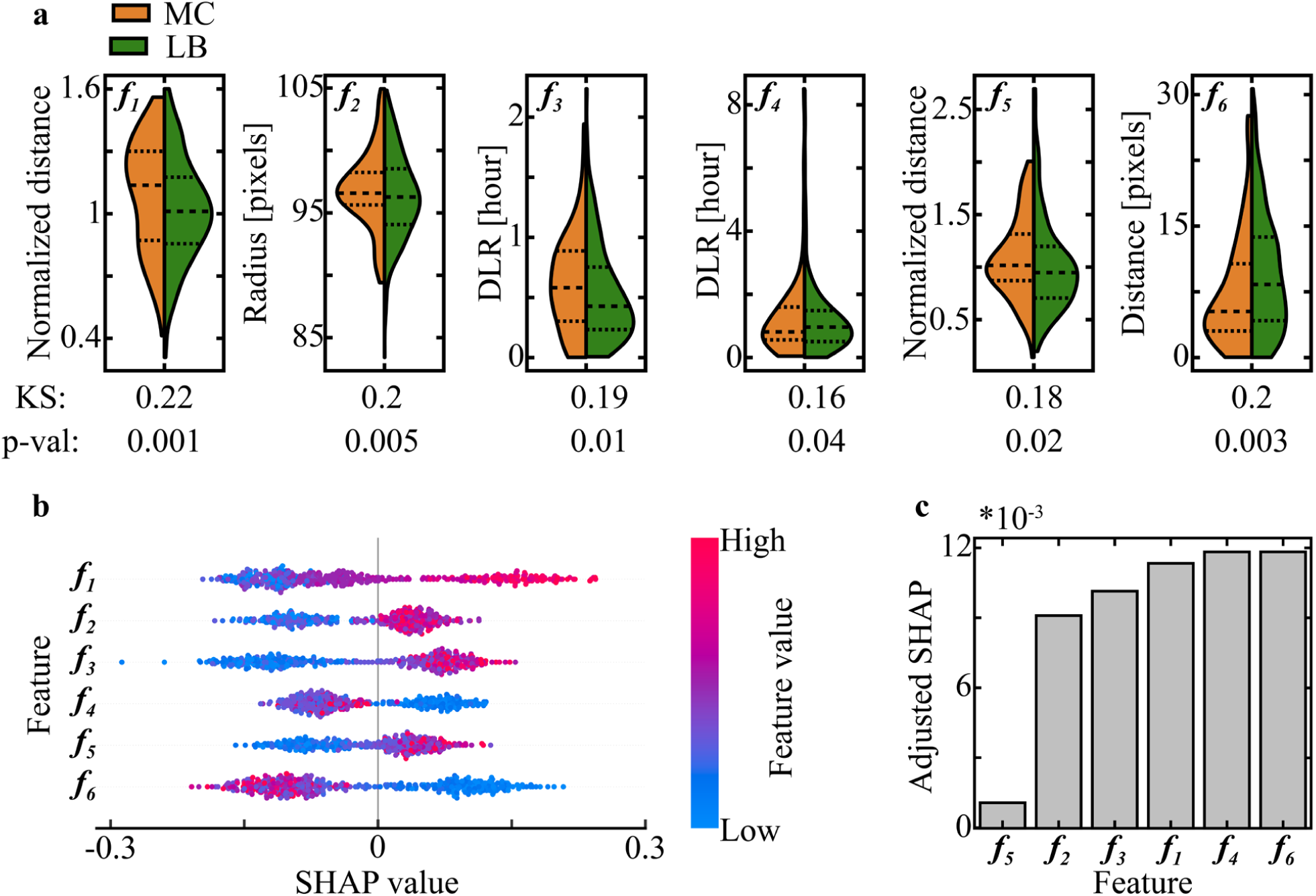
Statistical and SHAP feature analysis. **a**, Comparison of feature distributions shows small yet statistically significant differences between MC and LB embryos as measured by KS distances and p-value assessments. **b**, SHAP values are plotted versus feature values (color coded), showing a positive relationship between *f*_1_, *f*_2_, *f*_3_ and *f*_5_ and MC prediction, and a positive relationship between *f*_4_ and *f*_6_ and LB prediction. **c**, Feature importance is estimated by the aSHAP, which quantifies the impact on *accurate* prediction. KS: Kolmogorov-Smirnov. SHPA: Shapley Additive exPlantions. aSHAP: Adjusted SHAP.

To evaluate the impact that features have on MC prediction by the XG classifier, we employed the SHapley Additive exPlanations (SHAP) methodology.^44^ Features that obtained high positive SHAP values contribute to MC prediction of the embryos whereas high negative SHAP values contribute to LB prediction. We find that for all six features, the embryos with high feature values are separated from embryos with low features values along the SHAP axis (Fig. 4b). Specifically, high *f*_1_, *f*_2_, *f*_3_ and *f*_5_ values, which are obtained by embryos with positive SHAP, contribute to MC prediction. On the other hand, high *f*_4_ and *f*_6_ contribute to LB prediction (Fig. 4b). Unlike SHAP that measures the relationship between feature values and prediction, feature importance is assessed using the adjusted SHAP (aSHAP), which scores *accurate* prediction by adjusting a negative sign to false prediction.^20^ Indeed, we find that all the features of the final subset, with the exception of *f*_5_, hold comparable importance in directing accurate MC prediction (Fig. 4c).

## DISCUSSION

Non-invasive real-time evaluation of the risk of MC has the potential to improve embryo transfer protocols using time-lapse imaging. Training a machine learning-based classifier for assessing MC potential requires positively-implanted embryos. Hence, the available dataset is significantly smaller in size and morpho-dynamically less heterogeneous as compared with the prediction of embryo implantation potential. To overcome these limitations, we performed a rigorous feature screening and identified a subset of six morpho-dynamic features that retain prediction accuracy during early stages of preimplantation development. Interestingly, the selected subset was enriched with features that manifest the differences in the dynamic properties of PN and NPB compartments between MC and LB embryos. Since aneuploidy is tightly associated with first trimester MC,^5^ the contribution of PN and NPB-based features is consistent with a direct consequential impact that chromosomal abnormalities may have on PN and NPB dynamics. The fact that DLR2,3 and DLR5,6 features are included within the selected subset highlights potential downstream effects that chromosomal abnormalities may have on cleavage regulation in a manner that decouples early cleavage events (t2-t3) and late cleavage events (t5-t6). Importantly, feature screening as reported here is not deterministic and the selection of a different feature subset that is both predictive and non-redundant cannot be excluded. While the current features were extracted using semi-automated algorithms, clinical implementation would be further supported by developing fully-automated tools for the extraction of the reported features or of an equivalent subset of predictive and non-redundant features.

Time-lapse incubation systems provide high-quality visualization of preimplantation embryos that facilitate the development of machine learning-based classifiers. Such classifiers are routinely utilized within IVF clinics worldwide for selecting the embryos for transfer with the highest implantation potential. However, these algorithms are sensitive to various morphological elements and morphokinetic features that mark the potential to implant but fail to detect features that are associated with the risk of MC outcome. Using the same video files of preimplantation development, our algorithm predicts MC potential as early as the time of six-cells cleavage event (***f***_4_ : *DLR*_5,6_), thus supporting MC prediction within three days from fertilization in parallel to prediction of embryo implantation. Using our algorithm, assessing the risk of MC is compatible with assessing embryo implantation potential. The reported prediction of MC outcome with AUC 0.68% is comparable with the prediction accuracy of recently reported CNN implantation classifiers,^20,22^ suggesting that missing uterine parameters and/or other maternal factors prevent further improvement in the prediction of implantation outcome as well as MC outcome.^45^ In combination with implantation potential assessment, our algorithm thus provides a real-time non-invasive decision-support tool for deselecting embryos with high risk of MC outcome, which is expected to improve live-birth rates and shorten time-to-pregnancy in IVF-embryo transfer treatments.

## Data Availability

Due to ethical considerations and regulations by the Israeli Ministry of Health, no access is granted here to the data.

## ACKNOWLEDGMENT

We greatly appreciate support from the European Research Council (ERC-StG 678977). We thank Prof. Tommy Kaplan, Prof. Dafna Shahaf (Hebrew University of Jerusalem) and Dr. Assaf Ben-Meir (Hadassah Medical Center) for fruitful discussions. Author contributions: AB formulated and designed the research program. TA performed all computational analyses and was supported by YKT and NS. TA and AB designed and prepared the graphical material. AB wrote the manuscript. Competing interests: No financial or conflict of interests are declared.

## Supplementary Figures

**Figure S1:**
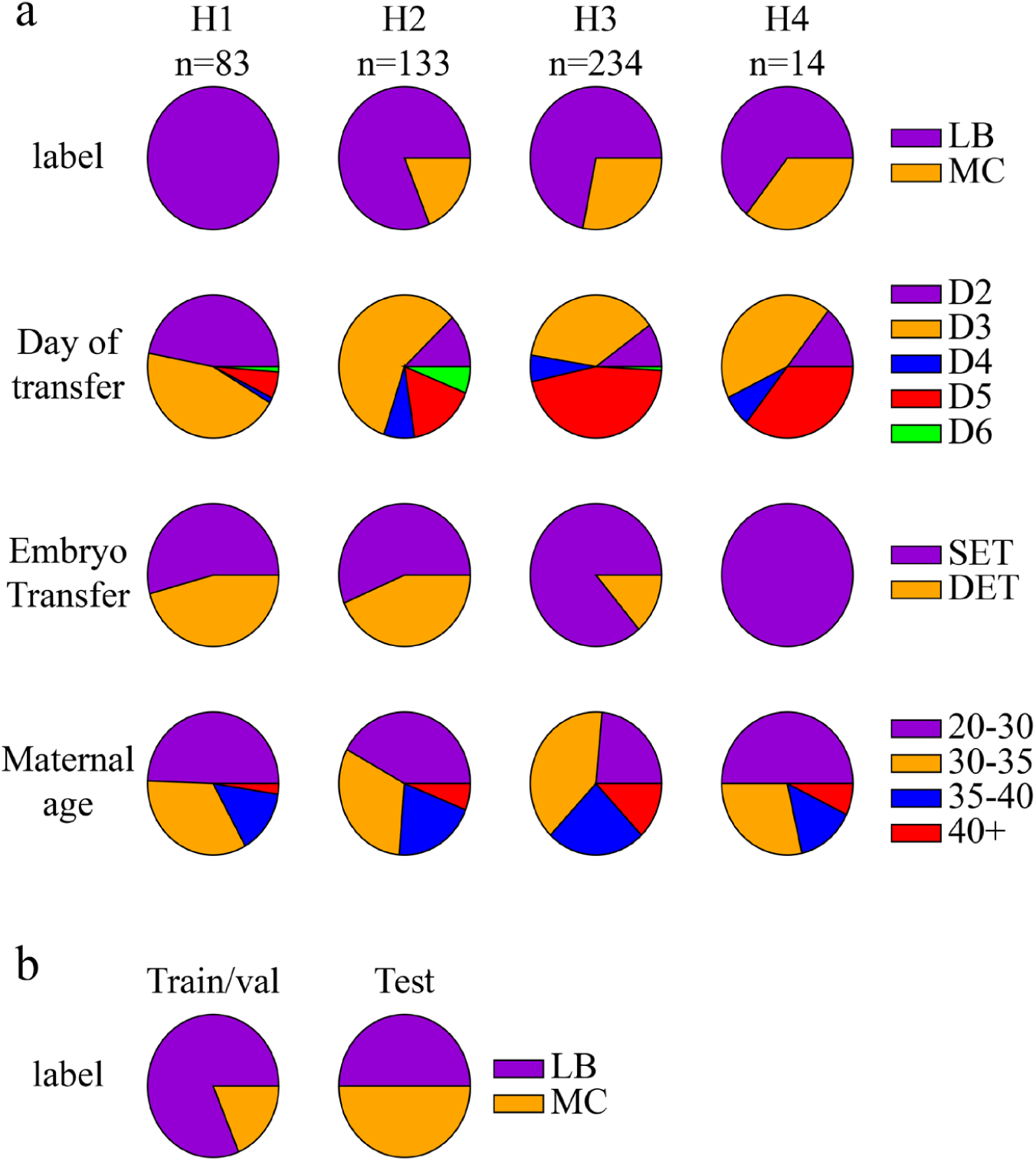
Characteristics of the dataset of 1^st^ trimester miscarriage (MC) and live birth (LB) embryos. **a;** Top to bottom: Distributions of MC/LB labelled embryos, day of transfer, number of transferred embryos and maternal age obtained from five data providing clinics (H1 to H5). **b;** The embryo dataset was divided into a train/validation set and an MC/LB-balanced test set.

**Figure S2:**
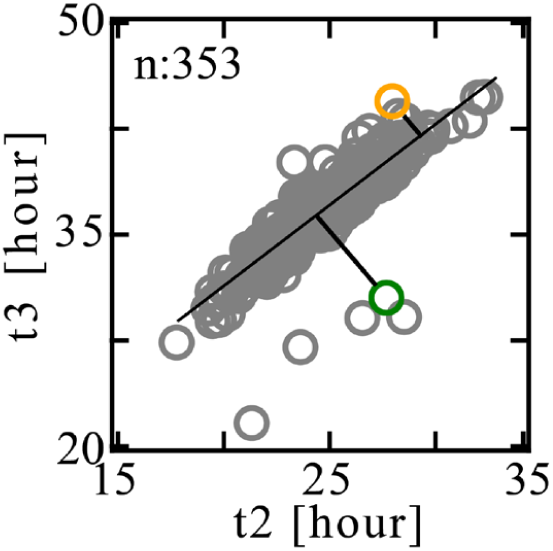
Distance from Linear Regression (DLR). Time of 3-cells cleavage (t3) is plotted versus time of 2-cells cleavage (t2) for a total of 353 LB embryos (gray symbols). The dependence between t3 and t2 is evaluated via linear regression (black line). t2-t3 DLR (DLR_2,3_) is the shortest distance of an embryo from the regression line as illustrated here for a short DLR_2,3_ embryo (marked in orange; 2.8 hours) and a high DLR_2,3_ embryo (marked in green; 6.4 hours).

**Figure S3:**
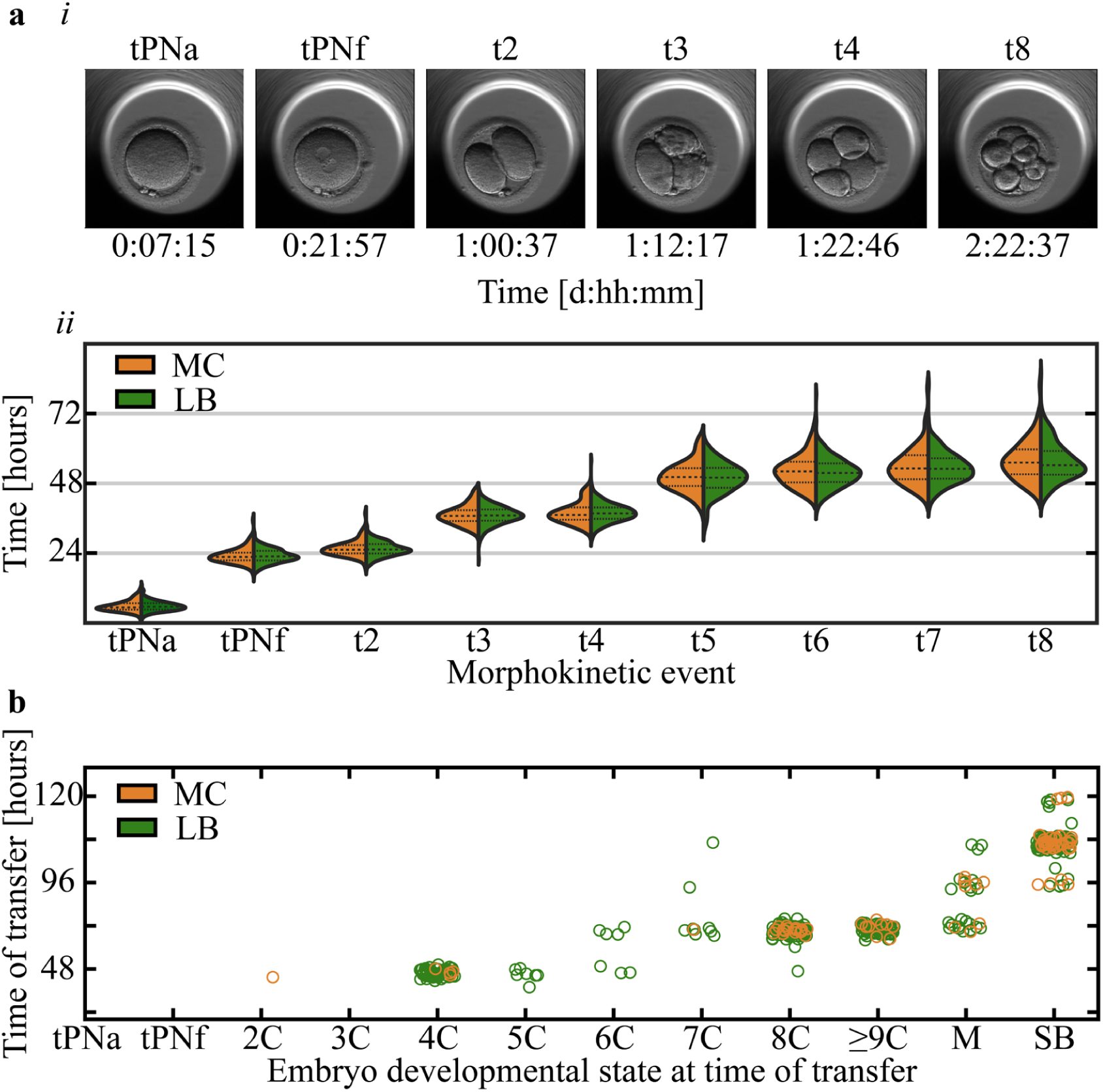
Live birth and 1st trimester miscarriage embryos share morphokinetic profiles, embryo state and time of transfer statistics. **a**, (i) Representative images of selected morphokinetic events from tPNa and tPNf to time of 8-cells cleavage (t8). (ii) Temporal distributions of LB and MC embryos are overlapping (KS p-values > 0.4). **b**, The relationship between time-of-transfer and embryo state at time-of-transfer is overlapping between MC and LB embryos. KS: Kolmogorov-Smirnov.

**Figure S4:**
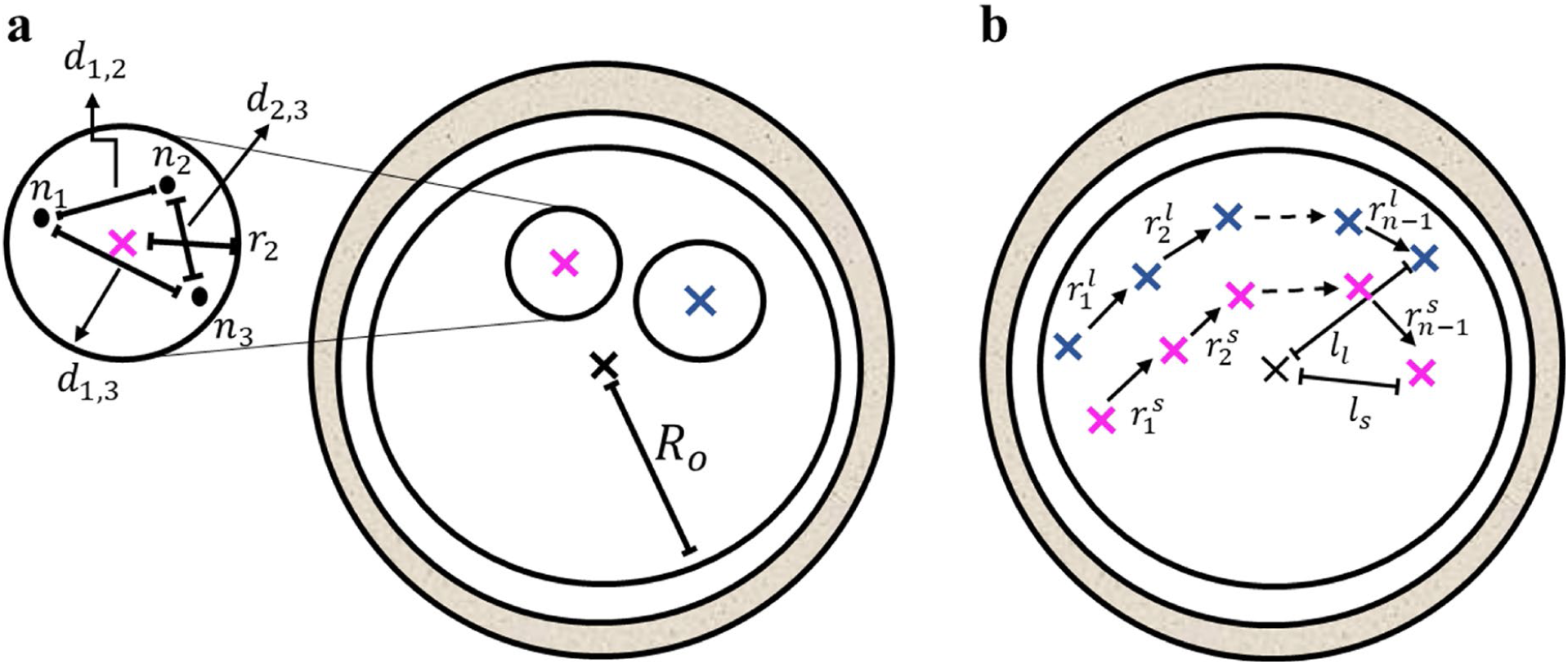
Illustration of the selected morpho-dynamic features for MC prediction. **a**, Feature *f*_1_ : The distances *d*_*ij*_ between all NPB pairs are zoomed-illustrated within the small PN. Feature *f*_2_ : The radius of the ooplasm, *R*_*o*_, and the center of mass of the ooplasm (black ‘×’) are depicted. Pink and blue ‘×’ marks represent the centers of mass of the small and large PN’i, respectively. **b**, Features *f*_5_ : *tPN*_*a*_ - to - *tPN*_*f*_ trajectories consist of (N-1) steps of the small (pink, 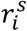) and large (blue, 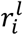) PN’i. Features *f*_6_ : The distances between the PN’i to the center of the zygote, *l*_*l*_ and *l*_*s*_, are evaluated at *tPN*_*f*_.

**Figure S5:**
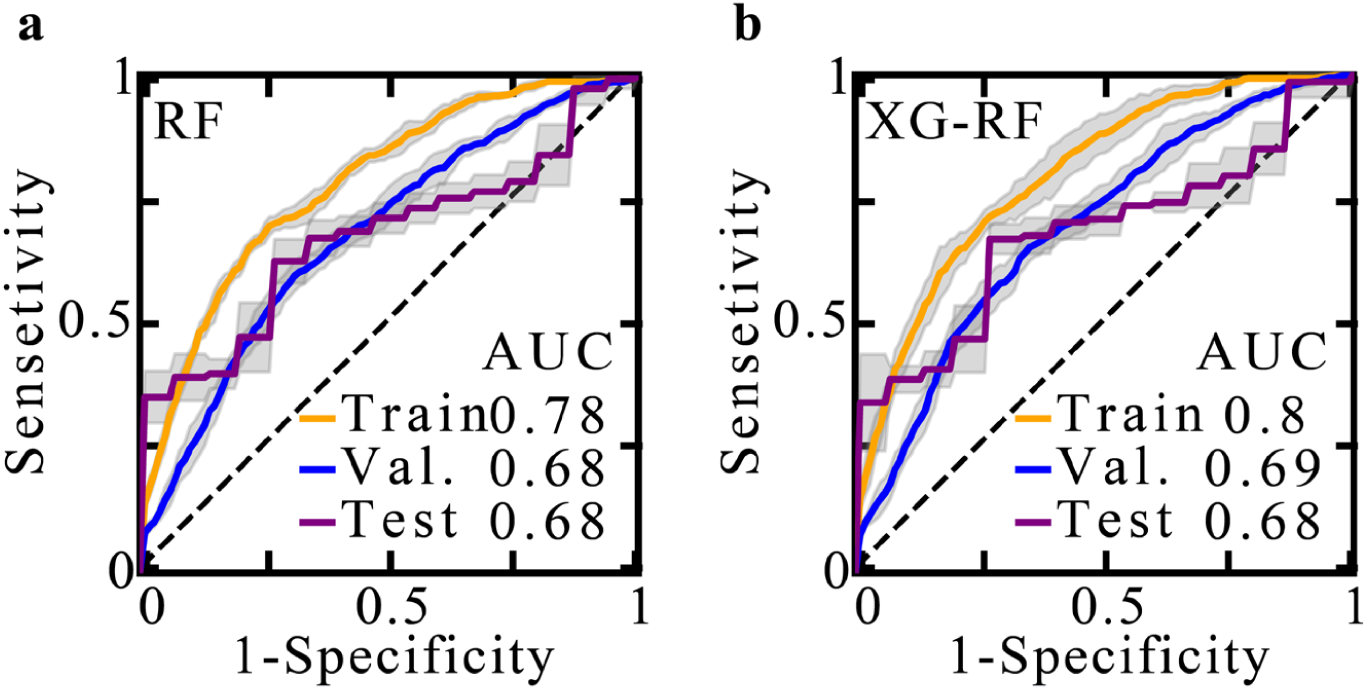
MC prediction by an RF and an RF-XG integrated classification models. Train, validation and test set ROC curves of MC prediction using an RF model and an integrated XG-RF classifier. The latter corresponds to averaging the MC scores that were obtained by RF and XG models. RF: Random forest. XG: XGBoost. ROC: Receiver operating characteristic. AUC: Area under the ROC curve.

